# High throughput wastewater SARS-CoV-2 detection enables forecasting of community infection dynamics in San Diego county

**DOI:** 10.1101/2020.11.16.20232900

**Authors:** Smruthi Karthikeyan, Nancy Ronquillo, Pedro Belda-Ferre, Destiny Alvarado, Tara Javidi, Christopher A. Longhurst, Rob Knight

## Abstract

Large-scale wastewater surveillance has the ability to greatly augment the tracking of infection dynamics especially in communities where the prevalence rates far exceed the testing capacity. However, current methods for viral detection in wastewater are severely lacking in terms of scaling up for high throughput. In the present study, we employed an automated magnetic-bead based concentration approach for viral detection in sewage that can effectively be scaled up for processing 24 samples in a single 40-minute run. The method compared favorably to conventionally used methods for viral wastewater concentrations with higher recovery efficiencies from input sample volumes as low as 10ml and can enable the processing of over 100 wastewater samples in a day. The sensitivity of the high-throughput protocol was shown to detect cases as low as 2 in a hospital building with a known COVID-19 caseload. Using the high throughput pipeline, samples from the influent stream of the primary wastewater treatment plant of San Diego county (serving 2.3 million residents) were processed for a period of 13 weeks. Wastewater estimates of SARS-CoV-2 viral genome copies in raw untreated wastewater correlated strongly with clinically reported cases by the county, and when used alongside past reported case numbers and temporal information in an autoregressive integrated moving average (ARIMA) model enabled prediction of new reported cases up to 3 weeks in advance. Taken together, the results show that the high-throughput surveillance could greatly ameliorate comprehensive community prevalence assessments by providing robust, rapid estimates.

**Importance:** Wastewater monitoring has a lot of potential for revealing COVID-19 outbreaks before they happen because the virus is found in the wastewater before people have clinical symptoms. However, application of wastewater-based surveillance has been limited by long processing times specifically at the concentration step. Here we introduce a much faster method of processing the samples, and show that its robustness by demonstrating direct comparisons with existing methods and showing that we can predict cases in San Diego by a week with excellent accuracy, and three weeks with fair accuracy, using city sewage. The automated viral concentration method will greatly alleviate the major bottleneck in wastewater processing by reducing the turnaround time during epidemics.

## Main

Wastewater-based epidemiology (WBE) can facilitate detailed mapping of the extent and spread of SARS-CoV-2 in a community and has seen a rapid rise in recent months owing to its cost effectiveness as well as its ability to foreshadow trends ahead of diagnostic testing (1-4). With over 46 million cases reported globally, 9.3 million of which are from the United States, there is an imminent need for rapid, community-level surveillance in order to identify potential outbreak clusters ahead of diagnostic data. Previous studies have reported high levels of correlation between viral concentration in sewage to clinically reported cases in a community with trends appearing 2-8 days ahead in wastewater (2, 5). A major bottleneck in large scale wastewater surveillance is the lack of robust, high throughput viral concentration methodology. Conventional techniques for viral concentration from wastewater typically employ laborious or time-consuming processes, namely polyethylene glycol (PEG) based precipitation, direct filtration or ultra-filtration methods that severely limit throughput (6). In the present study, we employed an affinity-capture magnetic hydrogel particle (Nanotrap) - based viral concentration method which was incorporated on the KingFisher Flex liquid-handling robot platform robots (Thermo Fisher Scientific, USA), using a 24-plex head to process 24 samples at once in a 40 min run. RNA is then extracted on the same Kingfisher system for rapid sample processing. Reverse transcription-quantitative polymerase chain reaction (RT-qPCR) targeting N1, N2, E-gene targets was used for detection and quantification of SARS-CoV-2 RNA. Detection of all 3 genes in a sample and its replicate (at Cq values <39) was considered positive. All steps from concentration to RT-qPCR plating were carried hands-free and carried out by liquid handlining robots to minimize human error. Using the above pipeline, 96 raw sewage samples were processed in a period of 4.5 hours (concentration to RT-qPCR detection/quantification) effectively reducing the processing time by at least 20-fold. The sensitivity of detection of SARS-CoV-2 viral RNA in wastewater by the high-throughput pipeline (i.e. to determine if individual buildings yield sufficient wastewater signal for high-resolution spatial studies) was established by routinely monitoring the SARS-CoV-2 signatures in the wastewater of a San Diego hospital building housing active COVID-19 patients (7). This site was used as a positive control to test correlations with caseload on a daily basis. Sewage samples were collected daily for a period of 12 weeks during which time the hospital’s case load (specific to Covid-19 patients) varied between 2-26. SARS-CoV-2 viral gene copies correlated with the daily hospital caseload (r=0.75, Suppl. Fig. S1) suggesting that the wastewater data could at least be used to identify the peaks. SARS-CoV-2 viral RNA was detected in wastewater on all days sampled. Furthermore, the high-throughput protocol has been used as a part of an on-campus wastewater surveillance for the last few months where data from 70 autosamplers covering individual buildings are sampled and analyzed on a daily basis. The method enabled detection of cases as low as 1 in large buildings (*manuscript in preparation, data available on request*).

The efficiency of high throughput concentration method was compared to the two other commonly implemented concentration methods (electronegative membrane filtration and Polyethylene glycol, PEG-based) using nine-fold serial dilutions of heat-inactivated SARS-CoV-2 viral particles spiked into 10 mL volumes of raw sewage which was previously verified to be from a location with no SARS-CoV-2 prevalence and verified by RT-qPCR (Suppl. Methods). The high-throughput protocol compared favorably to the conventionally used protocols demonstrating its potential implications for large scale sample processing (>100 samples/day) (Suppl. Fig. S2.A, Suppl. Fig. S3, Table S1). The average viral recovery efficiencies were 27% (SD 8%), 15% (SD 9%) and 13% (SD 8%) respectively for the high-throughput, PEG and HA filtration protocol (Suppl. Methods S1). The improved recovery could also be attributed in part due to the specificity of the magnetic beads in recovering viral particles coupled with a magnetic bead-based nucleic acid extraction protocol as well. The protocol was optimized on the KingFisher for optimal bead-binding through homogenization (dx.doi.org/10.17504/protocols.io.bptemnje). Furthermore, the automated protocol was hands-free and less prone to user discrepancies than the other 2 methods which were carried out manually.

24-hour flow weighted composites were collected each day for 3 months between July 20^th^-October 21^st^, 2020 from the influent stream of Point Loma wastewater treatment plant. The plant processes over 175 MGD (million gallons per day) of raw sewage and is the primary treatment center for the greater San Diego area serving over 2.3 million residents. SARS-CoV-2 viral RNA was detected in all of the samples processed at an average concentration of 2,010,104 gene copies/L in the influent. Over the course of the study, the clinically reported cases in the county increased by 29,375 (Fig. 1). Peaks in the wastewater data were frequently followed by peaks in the clinically confirmed cases at a later date. This suggests a correlation between wastewater and the number of new cases with the caveat of a time delay, where the wastewater data predicts future trends in the new number of cases. Although informative, this time-lagged correlation alone is not enough for robust predictions. This served as the main motivation to build a predictive model for forecasting the number of new cases per day in San Diego County. In order to model the number of new reported cases, which correlates, not only to wastewater but other complex prevalence and spreading dynamics, we embed useful predictors as separate time series to take advantage of any side information that may improve our forecasting results. Here, the day of the week was tracked for each data point in a third time series to capture any weekly trends for a total of three time series with 88 data points each. We used a data-driven approach to train a prediction model that utilizes prior reported cases, wastewater data and temporal correlations (embedded in the day of the week) in order to forecast the number of new positive cases in San Diego County. The multivariate Autoregressive Integrated Moving Average (ARIMA) model [7] was applied to build a prediction model for the number of new positive cases. The (predicted) number of new cases consist of lagged past values from all three series (number of new cases, wastewater data, day of the week) and each term can be thought of as the influence of that lagged time series on the number of new cases. 65 data points corresponding to data collected from 07/07/2020 to 09/28/2020, were used for developing the predictive model and forecasting estimates. The remaining 24 data points, corresponding to data collected from 09/29/2020 to 10/25/2020, were reserved for validating our forecast estimates. Fig. 2B shows the results of the model (prediction and forecast) compared to the observed data. The Pearson correlation coefficients and root mean squared error between the observed data and the predicted model were r = 0.84, 0.79, 0.69, 0.47, RMSE = 57, 50, 59, 70 for the trained model, 1, 2 and 3 week advance forecast values respectively (Suppl. Table S2). Our data-driven approach obtains forecasts successfully capturing general trends on the number of new cases (shown here up to 3 weeks in advance), ultimately reinforcing that wastewater analysis can be expository of previously undetected SARS-CoV-2 infections in the population which could provide useful insight for county officials for the purpose of early public health interventions. As with most environmental samples, there are biases and caveats associated with interpreting data from wastewater quantitatively, there are trade-offs while attempting to scale-up the process to enable the screening of hundreds of samples on a daily basis with a quick turn-around time. However, our data shows that we can still capture low prevalence cases enabling studies at high spatial resolution. Our study demonstrates that high-throughput wastewater-based surveillance can be successfully leveraged to enable creation of a rapid, large-scale early alert system for counties/districts and could be particularly useful in community surveillance in the more vulnerable populations.

**Figure 1:**
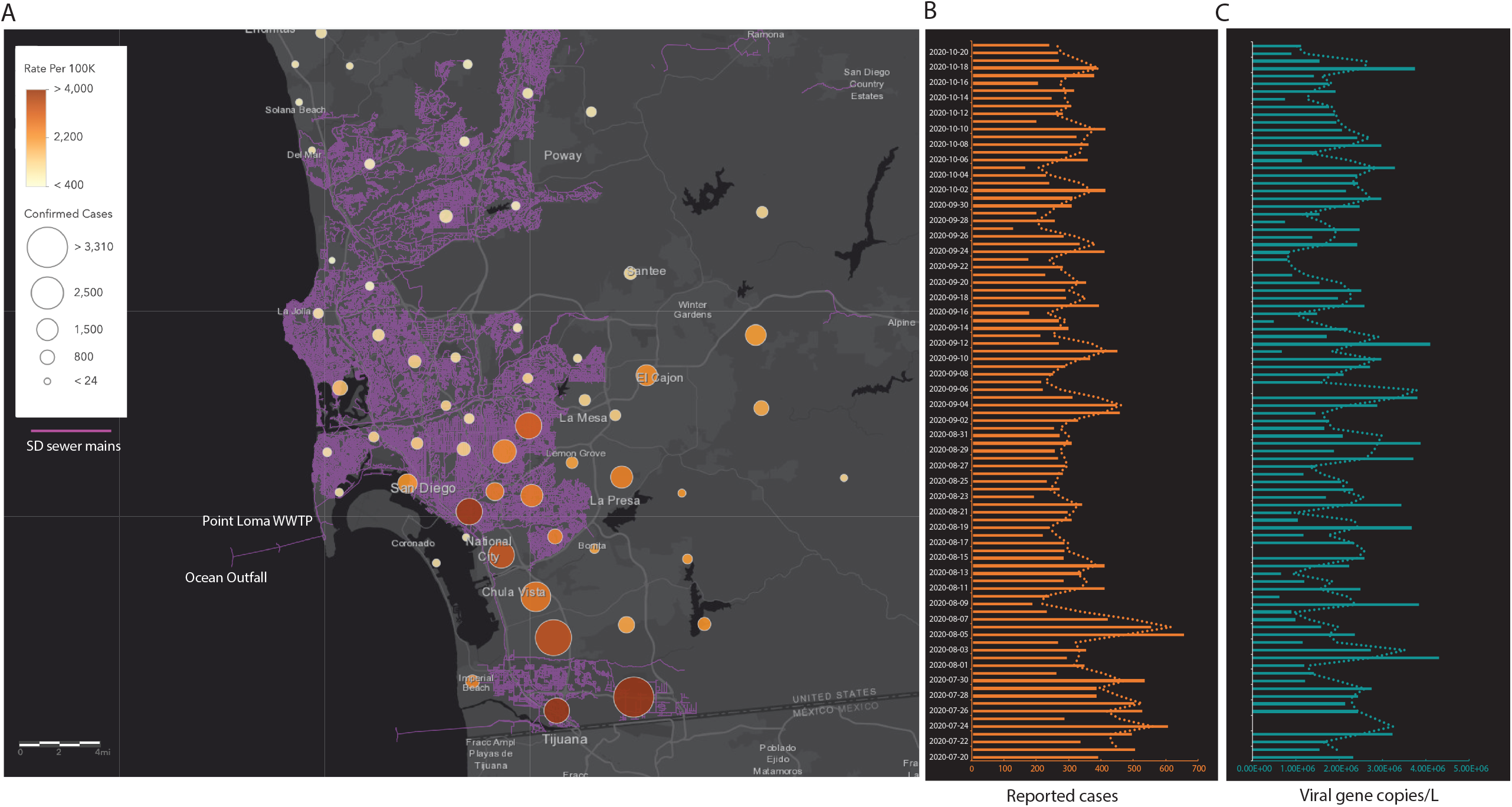
Tracking infection dynamics in SD county. **A**. Map showing the San Diego sewer mains (depicted in purple) that feed into the influent stream at the primary WWTP at Point Loma. Overlaid are the cumulative cases recorded from the different zip codes in the county during the course of the study. The caseload was counted by cases per zip code from areas draining into the WWTP. The circles are proportional to the diagnostic cases reported from each zone and the color gradient shows the cases per 100,000 residents. **B**. Daily new cases reported by the county of San Diego. **C**. SARS-CoV-2 viral gene copies detected per L of raw sewage determined from N1 Cq values corrected for PMMoV concentration (Pepper mild mottle virus). All viral concentration estimates were derived from the processing of 2 sample replicates and 2 PCR replicates for each sample.

**Figure 2:**
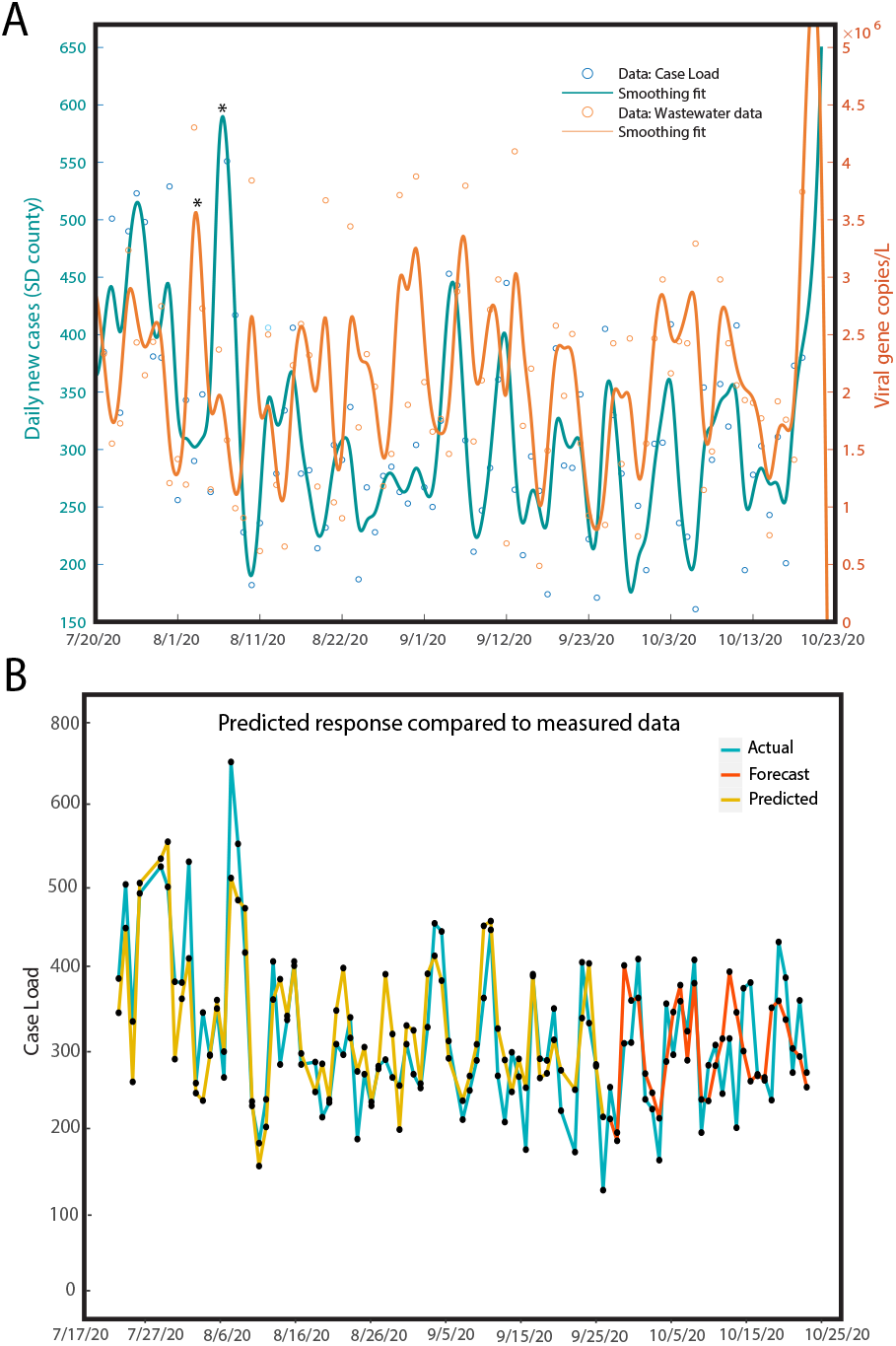
**A**. Daily caseload and wastewater viral concentration data shown for a period of 13 weeks, where a spline smoothing is applied to each time series to demonstrate general trends. **B**. Predictive model showing the predicted data (yellow) compared to the observed caseload (blue) and the 4-week forecast (red). Data collected from 07/07/2020 to 09/28/2020 were used as the training dataset to predict the caseload for the following weeks (up to 10/25/2020). Data (wastewater + county testing data) gathered from 09/29/2020-10/21/2020 were used for model validation. MATLAB Systems Identification toolbox was used to estimate the model order and parameters and calculate the forecasted values.

## Data Availability

All data referred to in the manuscript have been included as part of the manuscript or provided as supplemental files.

## ACKNOWLEDGEMENTS

The authors would like to thank the plant supervisor and the lab team at the Point Loma Wastewater Treatment plant in San Diego for providing us with samples and are grateful for their support in this effort. We thank Dr. Jack Gilbert and the Microbiome Sample Processing Core at UC San Diego for access to qPCR equipment. We also thank Jennifer Holland and the UC San Diego Health analytics team for generating the operational data for analysis. The authors would also like to thank Dr. Aaron Carlin from the Division of Infectious Diseases at UCSD for providing us with heat-inactivated SARS-CoV-2 viral particles. This work was supported by The University of California San Diego Return to Learn program (UCSD-RTL).

## Supplemental Data

**Fig. S1**: Plot showing the calculated viral gene copies/L of wastewater compared to the daily active COVID-19 caseload.

**Fig. S2**: A. Viral concentration method comparison. RT-qPCR N1 Cq values for nine-fold serial dilutions of heat-inactivated SARS-CoV-2 viral particles seeded into 10 mL volumes of raw sewage processed using the PEG concentration protocol (blue), Magnetic-bead based, Nanotrap protocol (purple) and electronegative membrane filtration method (gray). B,C,D. Standard curves for N1,N2 and E gene respectively for 6 fold-serial dilution of heat inactivated SARS-CoV-2 viral particles spiked into 10ml of raw sewage and concentrated using the high-throughput pipeline. Results for 3 replicates shown.

**Fig. S3**: Average N1 Cq values for 24 samples run with all 3 concentration methods.

**Fig. S4**: 1D Amplitude pictures of 2019nCOV ORF1a_ FAM

**Table S1**: N1 Cq values for the 24 samples concentrated with the 3 methods. SD (%) shows the standard deviation of the 2 replicates per sample run.

**Table S2**: Observed and predicted response values for the daily number of cases in San Diego county.

**Table S3**: A. Cycling conditions for RT-qPCR used for SARS-CoV-2 viral RNA detection using the multiplex Promega protocol. B. Cycling conditions for one-step ddPCR for viral quantification of the ORF1ab gene in the heat-inactivated SARS-CoV-2 viral particles used in recovery experiments. C. Dilution series of the heat-inactivated SARS-CoV-2 viral particles used in recovery experiments.

## Notes

### Competing Interest Statement

The authors have declared no competing interest.

### Clinical Protocols

https://www.protocols.io/view/automated-high-throughput-viral-concentration-from-bptemnje

