## Supplemental Data for "High throughput wastewater SARS-CoV-2 detection enables forecasting of community infection dynamics in San Diego county"

**Supplemental Methods**

**Wastewater viral concentration**

500 ml of 24-h flow weighted composite wastewater samples were collected from the Point Loma wastewater and transported on ice to the lab and stored at 4C until further processing. 5 samples were also collected from the effluent stream at ocean outfall. All samples were processed within 72h of sampling.

To test the sensitivity of detection, a commercial autosampler (HACH AS950) was deployed at the manhole immediately downstream of a San Diego hospital building to collect 24-h time-weighted composites (the sampler was programmed to collect samples every 30min throughout a 24h period for a duration of 12 weeks).

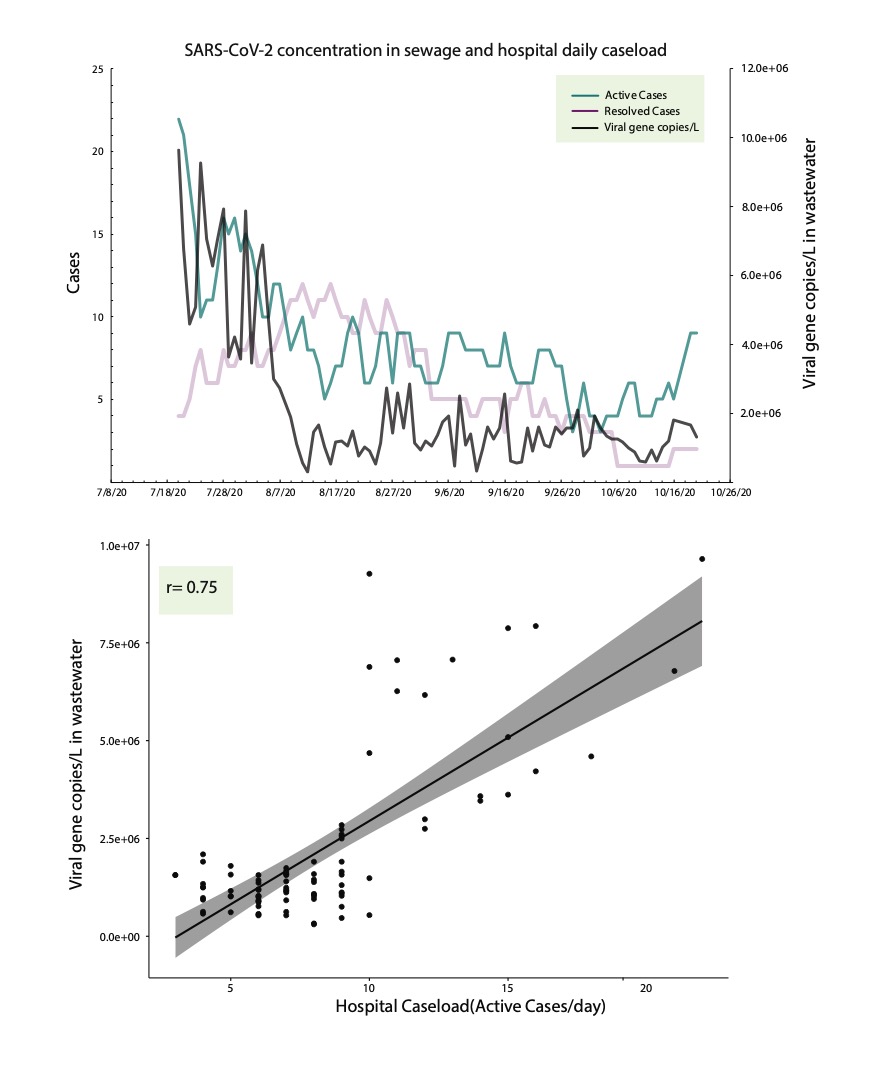

**Fig.S1: Plot showing the calculated viral gene copies/L of wastewater compared to the daily active COVID-19 caseload.**

*Magnetic bead based concentration:*

150 µL of affinity-capture magnetic hydrogel particles (Ceres Nanotrap) [1] were added to 10 ml of the well-mixed raw sewage sample (no pre-filtration step was carried out) in 24 deep well plates (Thermo Fisher Scientific, USA) and the viral concentration and elution step was performed on a KingFisher Flex robot (Thermo Fisher Scientific, USA). The concentrated viral particles were directly eluted in 150 µL of 1X PBS and 800 µL of viral lysis buffer of the MagMAX Microbiome Ultra Nucleic Acid Isolation Kit (Applied Biosystems, Cat # A24357, A24358). The downstream processing of the concentrated viral samples for nucleic acid extraction were carried out in 96 deep-well plates on the KingFisher Flex robot as per the manufacturer’s recommendations. Extracted nucleic acids were then eluted in 50 µL nuclease free water and used for SARS-CoV-2 real time RT-qPCR. Detailed protocol available at: **dx.doi.org/10.17504/protocols.io.bptemnje.**

*Electronegative membrane filtration:*

Samples were concentrated using 47 mm, 0.45 µm MCE filters using 500 mL magnetic filter funnels (Pall, cat # 28150-496) as described by Ahmed et al [2] without the addition of Mgcl_2_. Addition of Mgcl_2_ did not significantly alter the results when tested on a subset of the samples.

*Polyethylene glycol-8000 (PEG-8000) concentration:*

Samples were concentrated using PEG precipitation as described by Ahmed et al [3] (Method F, with the slight modification where the samples were incubated overnight at 4C instead of 2h).

The nucleic acid extraction and RT-qPCR steps were identical for all 3 viral concentration methods.

**RT-qPCR detection of SARS-CoV-2 viral RNA**

SARS-CoV-2 viral RNA detection and quantification were carried out using the Center for Disease Control (CDC) 2019-Novel Coronavirus Real-Time RT-PCR Diagnostic Panel and the E-gene primer/probe from the World Health Organization [4]. The reactions were carried out in a 384 well plate using a miniaturized version of the CDC protocol. Each sample underwent 4 separate RT-qPCR reactions; 3 reactions targeting specifically SARS-CoV-2 (N1 and N2 target the nucleocapsid and the E-gene targets a virporin forming gene) and 1 reaction targeting a human gene (RNAse P gene) which served as a positive control for sample collection and nucleic acid extraction. 4µL of the template was used for a total of 10µL reaction mix which also consisted of 2µL of RNAse-free water, 1µL of primer/probe mix and 3uL of TaqPath™ 1-Step RT-qPCR Master Mix (Life Sciences, Thermo Fisher Scientific, USA). Sample plating was performed using an EpMotion automated liquid handler (Eppendorf, Germany). 2 positive controls and 2 extraction blanks were included in every run. The RT-qPCR reactions were carried out in a CFX384 Real-Time System (BIO-RAD) thermocycler (cycling conditions shown below).

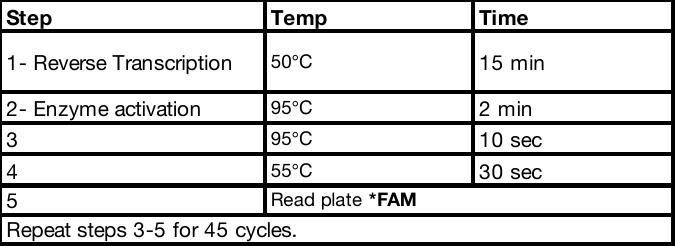

*Inhibition and extraction control*

Presence of RT-qPCR reaction inhibitors are commonly encountered in wastewater matrices (especially those that have a higher degree of solids) and in order to screen for inhibitors, all the RT-qPCR reactions were also run using Promega SARS-CoV-2 RT-qPCR Kit for

Wastewater (Cat.no. CS317402, Promega, USA). Primer/probe sets targeting the N1, N2, E gene; primers detecting Pepper Mild Mottle Virus (PMMoV) as an internal process control; and an internal amplification control, IAC (for inhibition assessment) were used. The samples were re-run for the RT-qPCR step in order to estimate the PMMoV values (to assess significant changes in PMMoV values that could account for anomalies in the wastewater peaks). For all samples processed, IAC Cq values were within the acceptable range of 20-25 (mean 21, SD 0.2) and PMMoV Cq values were between 22-24. SARS-CoV-2 RNA were quantified as viral gene copies/L of raw sewage by applying a linear regression to the standard curve comprising of nine-fold serial dilution of heat-inactivated SARS-CoV-2 viral particles and calculating the best fit of the standard curve using y = mx + b, where x = log concentration, y = Cq, and m=slope (where m= –3.3 indicates 100% PCR efficiency). Furthermore, to detect inhibition specific to sewage samples, the positive control RNA ladder was run with every qPCR run (5 1:10 serial dilutions of a positive control). We did not find any significant differences for dilutions in nuclease free-water and the sewage extracts (verified to be SARS-CoV-2 negative) spiked in with the same dilutions. Cq vales for the no dilution to 1:100000 dilution were not significantly different suggesting no PCR inhibition in the RNA extracts. Two-tailed t tests at 95% confidence interval was used to determine if the average Cq values were statistically significant from the spiked-in water control.

The RT-qPCR reactions were carried out in a CFX384 Real-Time System (BIO-RAD) thermocycler (cycling conditions shown below).

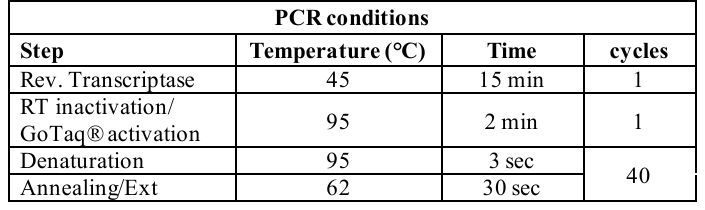

3 biological replicates were performed per dilution. Inactivated viral particles (obtained from the UCSD CFAR BSL3 facility) were quantified using the ORF1ab gene. The viral particles were quantified using 1-step ddPCR (Suppl. Table S3, Suppl. Fig. S4).

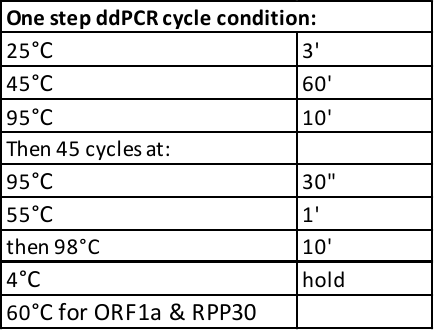

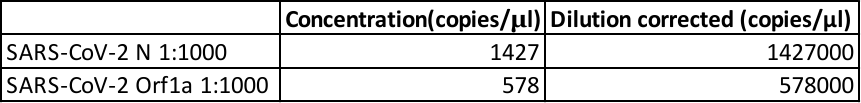

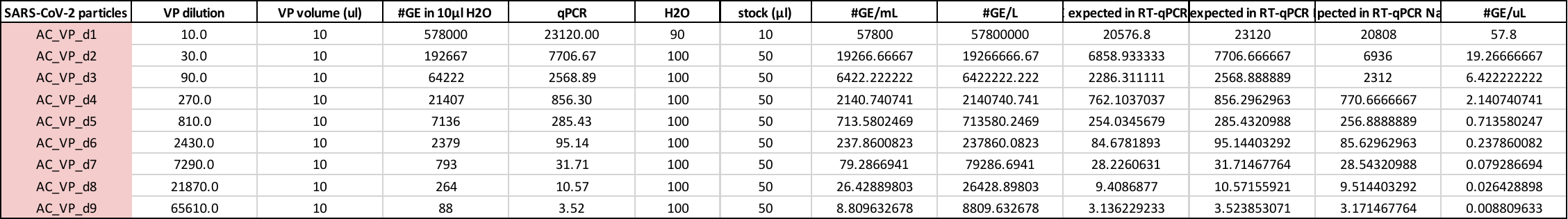

The 1D Amplitude pictures of 2019nCOV ORF1a_ FAM:

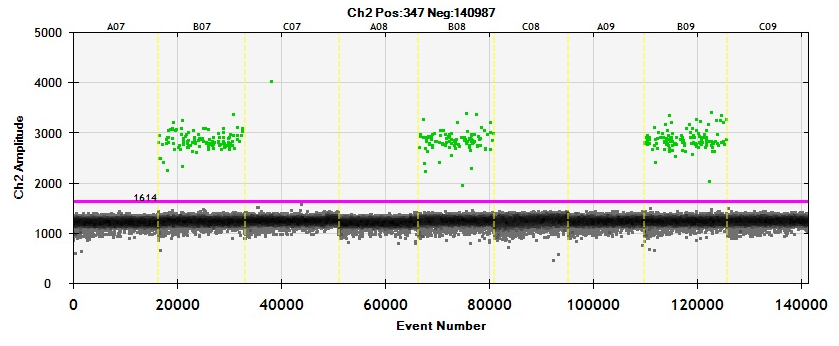

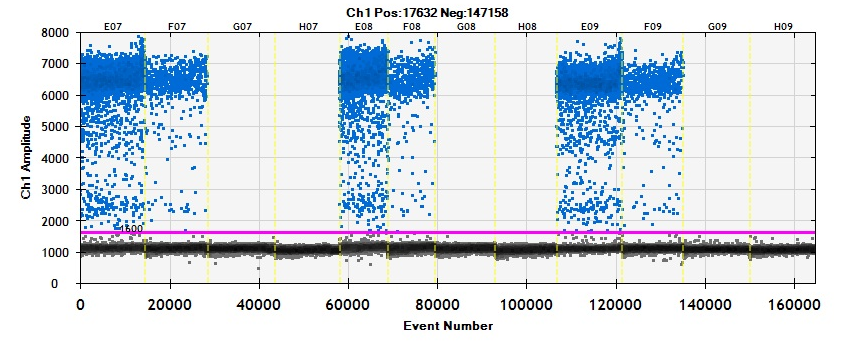

The SARS-CoV-2 recovery efficiency of each viral concentration method for each replicate was calculated by the equation:

$$\% Recovery=\frac{viral RNA GE recovered}{viral RNA GE seeded}\times100$$

Viral RNA GE recovered was evaluated with the standard curve generated from the direct RT-qPCR of the quantified SARS-CoV-2 viral particle dilution series.

Mean and standard deviation % recovery of SARS-CoV-2 for nine standard dilutions per all viral concentration methods.

| Concentration Methods | Mean | Standard Deviation |
| --- | --- | --- |
| Nanotrap | 23% | 8% |
| PEG | 15% | 9% |
| HA Filter | 13% | 8% |

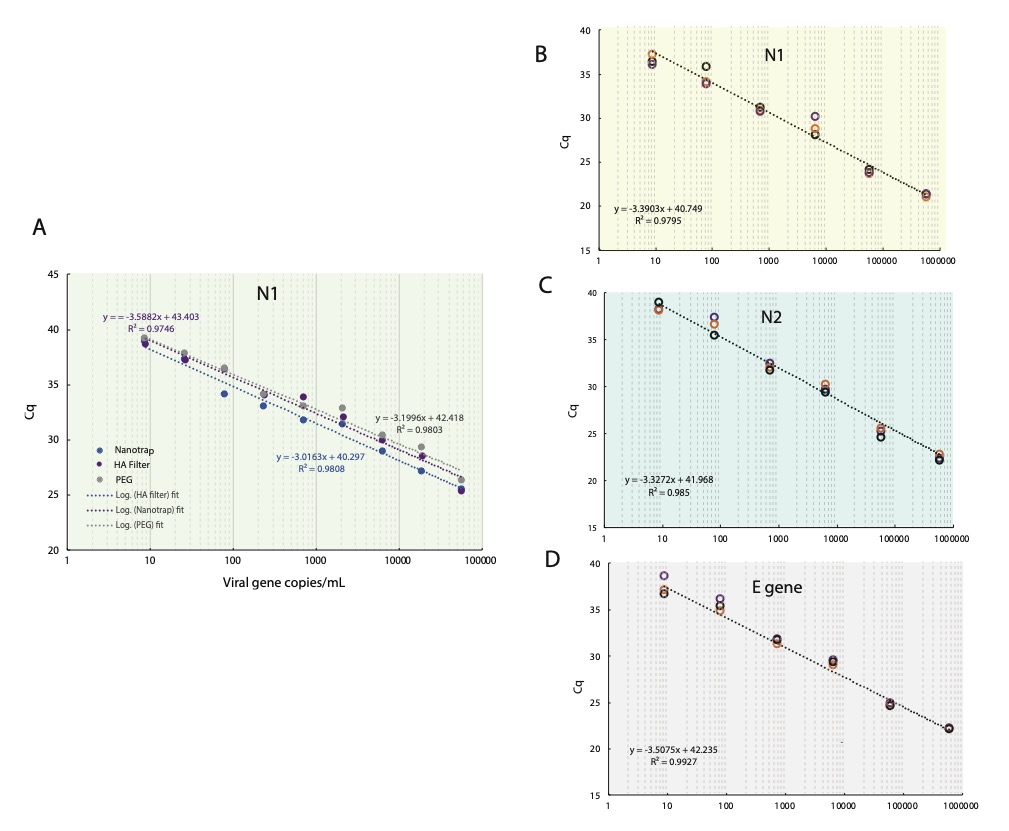

**Suppl. Fig. S2: A.** **Viral concentration method comparison**. RT-qPCR N1 Cq values for nine-fold serial dilutions of heat-inactivated SARS-CoV-2 viral particles seeded into 10 mL volumes of raw sewage processed using the PEG concentration protocol (blue), Magnetic-bead based, Nanotrap protocol (purple) and electronegative membrane filtration method (gray). B,C,D. Standard curves for N1,N2 and E gene respectively for 6 fold-serial dilution of heat-inactivated SARS-CoV-2 viral particles spiked into 10ml of raw sewage and concentrated using the high-throughput pipeline. Results for 3 replicates shown.

The level of suspended solids can greatly differ depending on the type of wastewater, consequently affecting the recoveries due to viral adsorption to solids.

In order to test the efficacy of the high throughput method on actual samples, a subset of 24 samples were run through all 3 concentration protocols (downstream processing kept identical). The N1 Cq values and their SD are shown in Suppl. Table S1 and Suppl. Fig. S3.

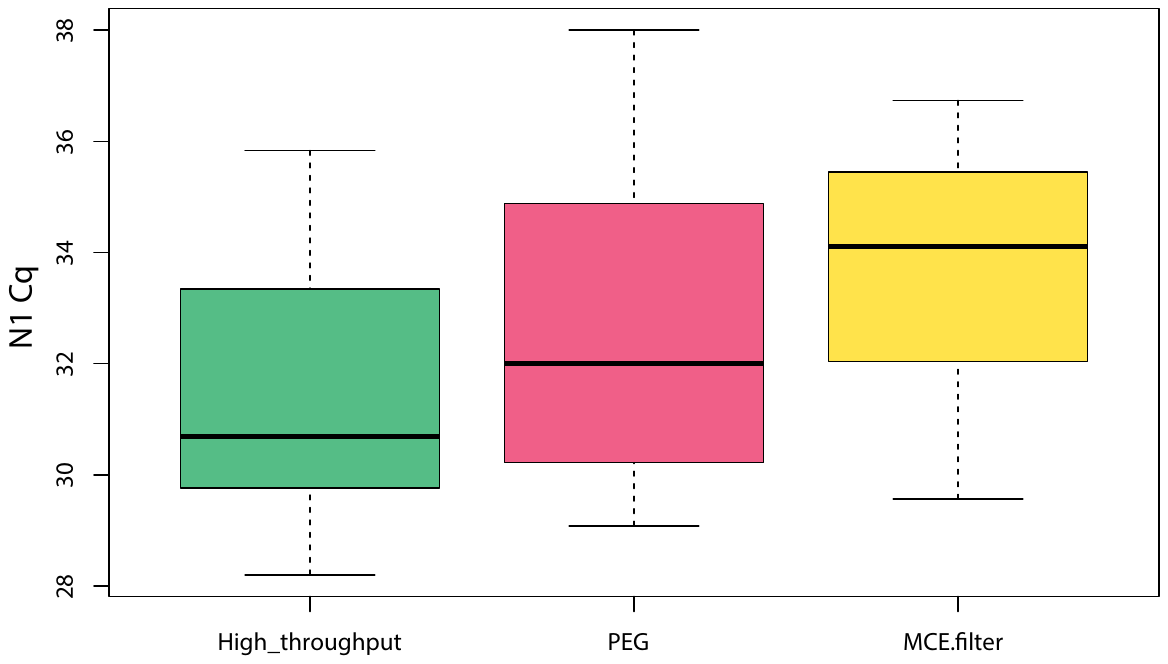

**Fig.S3: Average N1 Cq values for 24 samples run with all 3 concentration methods.**

N1 was primarily used in quantification as it has been shown to be more reliable/sensitive for detection [5].

**Table S1: N1 Cq values for the 24 samples concentrated with the 3 methods. SD (%) shows the standard deviation of the 2 replicates per sample run.**

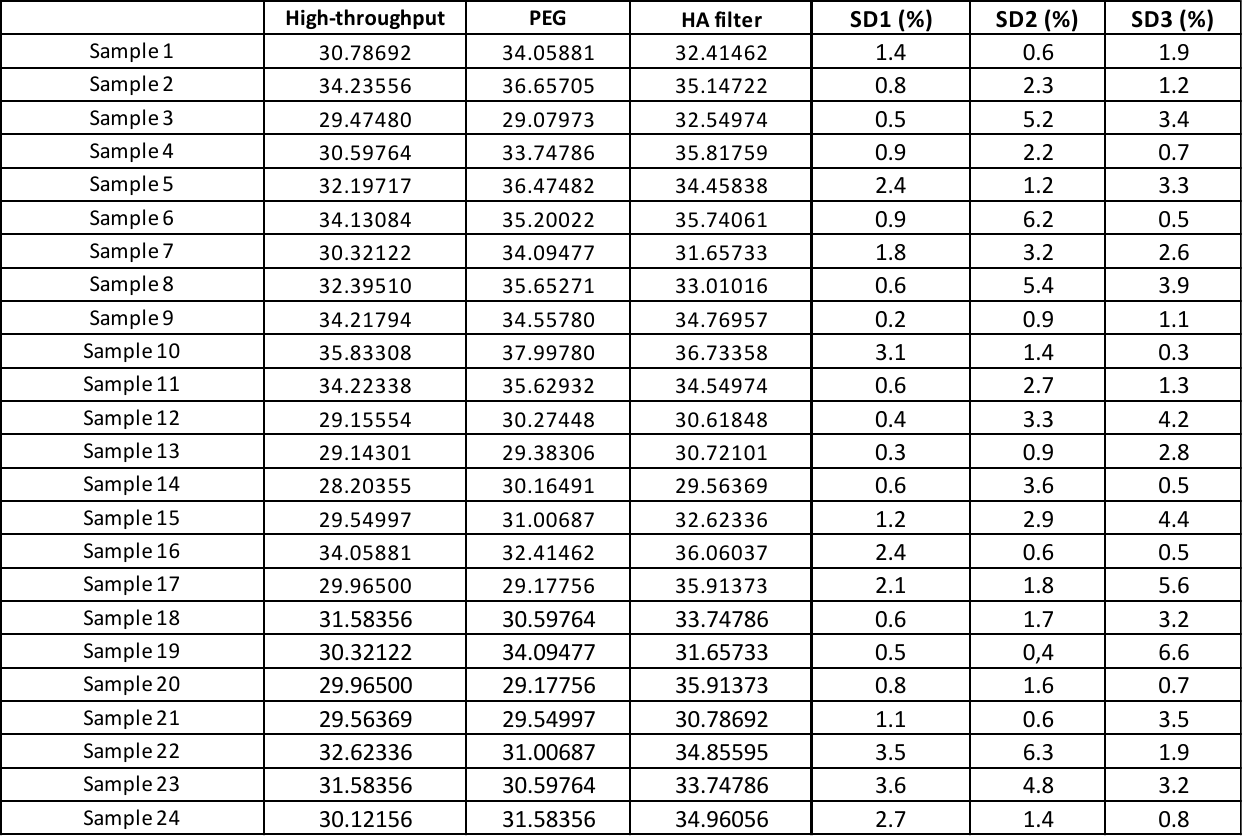

**Table S2: Observed and predicted response values for the daily number of cases in San Diego county**

|  | **Daily new cases (SD county)** | | | **Viral gene copies/L** | **Day of the week** |
| --- | --- | --- | --- | --- | --- |
| **Date** | Data | Estimated Model | Forecast values | Wastewater data | 1=Monday |
| 7/20/20 | 385 | 342.7569 |  | 2,332,026.54 | 1 |
| 7/21/20 | 501 | 447.1958 |  | 1,551,857.02 | 2 |
| 7/22/20 | 332 | 257.5234 |  | 1,727,150.62 | 3 |
| 7/23/20 | 490 | 502.6874 |  | 3,235,187.12 | 4 |
| 7/26/20 | 523 | 532.747 |  | 2,431,730.76 | 7 |
| 7/27/20 | 498 | 553.6315 |  | 2,146,578.31 | 1 |
| 7/28/20 | 381 | 285.5421 |  | 2,437,051.95 | 2 |
| 7/29/20 | 380 | 360.0118 |  | 2,747,828.41 | 3 |
| 7/30/20 | 529 | 409.7799 |  | 1,207,764.74 | 4 |
| 7/31/20 | 256 | 243.7113 |  | 1,417,154.42 | 5 |
| 8/1/20 | 343 | 234.4321 |  | 1,194,551.65 | 6 |
| 8/2/20 | 290 | 290.8771 |  | 4,303,251.63 | 7 |
| 8/3/20 | 348 | 358.3898 |  | 2,728,333.35 | 1 |
| 8/4/20 | 263 | 294.8408 |  | 1,152,773.68 | 2 |
| 8/5/20 | 652 | 508.8962 |  | 2,369,773.37 | 3 |
| 8/6/20 | 551 | 481.5993 |  | 1,582,051.11 | 4 |
| 8/7/20 | 417 | 471.6133 |  | 988,905.61 | 5 |
| 8/8/20 | 228 | 233.411 |  | 904,293.88 | 6 |
| 8/9/20 | 182 | 153.7541 |  | 3,842,016.87 | 7 |
| 8/10/20 | 236 | 201.9887 |  | 616,073.46 | 1 |
| 8/11/20 | 406 | 359.0887 |  | 2,500,428.42 | 2 |
| 8/12/20 | 279 | 384.1577 |  | 1,192,246.95 | 3 |
| 8/13/20 | 334 | 338.8829 |  | 655,394.59 | 4 |
| 8/14/20 | 406 | 400.727 |  | 2,233,170.30 | 5 |
| 8/15/20 | 279 | 292.5592 |  | 2,592,742.60 | 6 |
| 8/17/20 | 282 | 245.1322 |  | 2,320,963.19 | 1 |
| 8/18/20 | 214 | 279.8132 |  | 1,177,421.34 | 2 |
| 8/19/20 | 232 | 235.8228 |  | 3,669,143.04 | 3 |
| 8/20/20 | 304 | 345.5726 |  | 1,040,025.39 | 4 |
| 8/21/20 | 291 | 397.916 |  | 901,987.27 | 5 |
| 8/22/20 | 337 | 311.7353 |  | 3,442,221.79 | 6 |
| 8/23/20 | 187 | 270.4436 |  | 1,692,578.54 | 7 |
| 8/24/20 | 267 | 300.3693 |  | 2,330,169.78 | 1 |
| 8/25/20 | 228 | 232.7673 |  | 2,046,962.79 | 2 |
| 8/26/20 | 277 | 273.5418 |  | 1,180,550.83 | 3 |
| 8/27/20 | 285 | 390.2893 |  | 1,461,755.50 | 4 |
| 8/28/20 | 263 | 316.5342 |  | 3,714,923.59 | 5 |
| 8/29/20 | 253 | 198.707 |  | 1,888,830.98 | 6 |
| 8/30/20 | 304 | 327.0192 |  | 3,877,063.78 | 7 |
| 8/31/20 | 267 | 321.109 |  | 2,086,813.19 | 1 |
| 9/1/20 | 250 | 255.4555 |  | 1,655,486.25 | 2 |
| 9/2/20 | 325 | 390.839 |  | 1,767,155.04 | 3 |
| 9/3/20 | 453 | 412.9572 |  | 1,461,755.50 | 4 |
| 9/4/20 | 443 | 382.3747 |  | 2,875,872.26 | 5 |
| 9/5/20 | 308 | 286.561 |  | 3,796,334.60 | 6 |
| 9/7/20 | 211 | 234.3069 |  | 1,570,126.24 | 1 |
| 9/8/20 | 247 | 264.8406 |  | 2,102,392.59 | 2 |
| 9/9/20 | 284 | 303.8505 |  | 2,716,640.54 | 3 |
| 9/10/20 | 361 | 449.9258 |  | 2,980,097.51 | 4 |
| 9/11/20 | 445 | 455.8139 |  | 682,574.24 | 5 |
| 9/12/20 | 265 | 323.2479 |  | 4,095,336.32 | 6 |
| 9/13/20 | 208 | 284.5112 |  | 1,705,893.20 | 7 |
| 9/14/20 | 294 | 245.2321 |  | 2,204,297.45 | 1 |
| 9/15/20 | 264 | 285.8503 |  | 487,224.07 | 2 |
| 9/16/20 | 174 | 250.3876 |  | 1,490,048.76 | 3 |
| 9/17/20 | 388 | 390.2926 |  | 2,578,090.07 | 4 |
| 9/18/20 | 286 | 262.3267 |  | 1,966,959.29 | 5 |
| 9/19/20 | 284 | 268.0696 |  | 2,505,701.77 | 6 |
| 9/20/20 | 348 | 309.3863 |  | 1,552,473.50 | 7 |
| 9/21/20 | 222 | 271.8353 |  | 925,178.92 | 1 |
| 9/23/20 | 171 | 247.8889 |  | 816,590.99 | 3 |
| 9/24/20 | 405 | 336.211 |  | 843,455.66 | 4 |
| 9/25/20 | 330 | 403.6176 |  | 2,424,640.19 | 5 |
| 9/26/20 | 279 | 276.8502 |  | 1,373,821.46 | 6 |
| 9/27/20 | 124 | 214.3101 |  | 2,466,391.42 | 7 |
| 9/28/20 | 251 |  | 211.8419 | 743,916.77 | 1 |
| 9/29/20 | 195 |  | 184.8794 | 1,552,462.51 | 2 |
| 9/30/20 | 305 |  | 401.1089 | 2,466,391.42 | 3 |
| 10/1/20 | 306 |  | 357.901 | 2,980,097.51 | 4 |
| 10/2/20 | 409 |  | 361.1006 | 2,163,129.52 | 5 |
| 10/3/20 | 236 |  | 267.7269 | 2,441,459.52 | 6 |
| 10/4/20 | 224 |  | 244.0649 | 2,424,640.20 | 7 |
| 10/5/20 | 161 |  | 212.6933 | 3,292,463.35 | 1 |
| 10/6/20 | 354 |  | 282.607 | 1,147,833.24 | 2 |
| 10/7/20 | 291 |  | 343.6412 | 1,483,191.26 | 3 |
| 10/8/20 | 357 |  | 376.8217 | 2,980,097.51 | 4 |
| 10/9/20 | 320 |  | 379.2427 | 2,424,640.19 | 5 |
| 10/10/20 | 408 |  | 284.1227 | 2,059,575.59 | 6 |
| 10/11/20 | 195 |  | 235.8501 | 1,930,223.93 | 7 |
| 10/12/20 | 278 |  | 234.0864 | 1,907,001.33 | 1 |
| 10/13/20 | 303 |  | 277.6383 | 1,771,271.45 | 2 |
| 10/14/20 | 243 |  | 310.8178 | 753,837.19 | 3 |
| 10/15/20 | 311 |  | 393.3864 | 1,919,260.63 | 4 |
| 10/16/20 | 201 |  | 343.2658 | 1,759,581.55 | 5 |
| 10/17/20 | 373 |  | 295.4904 | 1,411,080.92 | 6 |
| 10/18/20 | 380 |  | 258.3931 | 3,742,657.88 | 7 |
| 10/19/20 | 265 |  | 266.7648 | 1,553,446.52 | 1 |
| 10/20/20 | 263 |  | 259.4196 | 909,420.24 | 2 |
| 10/21/20 | 235 |  | 348.9845 | 1,115,330.21 | 3 |
| 10/22/20 | 430 |  | 357.4664 |  | 4 |
| 10/23/20 | 386 |  | 334.3438 |  | 5 |
| 10/24/20 | 269 |  | 298.7001 |  | 6 |
| 10/25/20 | 358 |  | 288.7466 |  | 7 |
| 10/26/20 | 269 |  | 251.0131 |  | 1 |
